# A Digital Environment for Multi-modal Relational Management, Representation, and Analysis of Cancer Clinical Studies

**DOI:** 10.64898/2026.10.01.26364400

**Authors:** Ferran Cardoso Rodriguez, Xixuan Zhu, Eugene F. Schuster, Lucy Kilburn, Holly Tovey, Aude Espinasse, Ruth Sardinha, Tom Lund, Alistair Ring, Manuel Salto-Tellez, Maggie Chon U Cheang

## Abstract

Biologically rich clinical trials generate heterogeneous datasets whose relationships and clinical context must be preserved. Here we present the POETIC-AI environment, a trial-centric digital environment for integrating, analysing, and exploring multimodal data from POETIC, a randomised phase III trial of peri-operative aromatase inhibition in hormone receptor-positive breast cancer (NCT02338310).

We implemented a graph-based ontology linking patients, treatments, sampling timepoints, specimens, assays, images, spatial regions, and derived measurements; and designed code-based and graphical analytical workflows. Using proliferation as a test case, we reproduced POETIC’s established reduction in Ki67% and examined variation underlying sample-level measurements. In response-enriched cohorts, mean Ki67% decreased while relative variation among pathologist-assessed fields increased (paired Wilcoxon, P < 0.0001). Pathologist-assessed Ki67% correlated strongly with an 18-gene transcriptomic proliferation signature (*r* = 0.834), and both decreased significantly during treatment (P < 0.0001). An annotation-free pixel-wise Ki67 H-score showed weaker correlations and a similar but non-significant reduction (*r* = 0.327-0.472; P = 0.15), highlighting differences between tumour-focused and whole-section image quantification. Digital pathology and spatial molecular measurements remained linked to their source images and annotated tissue regions, enabling exploration within their morphological and spatial context using bespoke interactive applications.

POETIC-AI demonstrates how the structure, provenance, and tissue context of evolving multimodal clinical-trial data can be preserved within a connected research environment, supporting reproducible analyses and the laying the groundwork for future development of biologically interpretable computational biomarkers.

## Introduction

Clinical trials with biologically rich translational research programmes increasingly generate heterogeneous data across clinical, molecular, imaging, and spatial modalities. Integrating these data provides opportunities to characterise treatment response across biological modalities and to develop artificial intelligence (AI) approaches that identify relationships within and across modalities. However, this requires data generated using very different technologies to remain connected to their clinical and biological contexts.

Peri-Operative Endocrine Therapy—Individualising Care (POETIC, NCT02338310) was a randomised phase 3 clinical trial evaluating peri-operative aromatase inhibitor (POAI) therapy in hormone receptor-positive early breast cancer. Patients were randomly assigned 2:1 ratio to POAI or control groups, with those allocated to the POAI group receiving two weeks of aromatase inhibitor before and after surgery (Figure 1A). Although POAI did not improve long-term clinical outcomes, POETIC demonstrated the prognostic value of measuring tumour proliferation via Ki67 immunohistochemistry (IHC) at baseline and during treatment; with on-treatment Ki67% predicting long-term outcome in patients with elevated baseline proliferation^1^.

**Figure 1:**
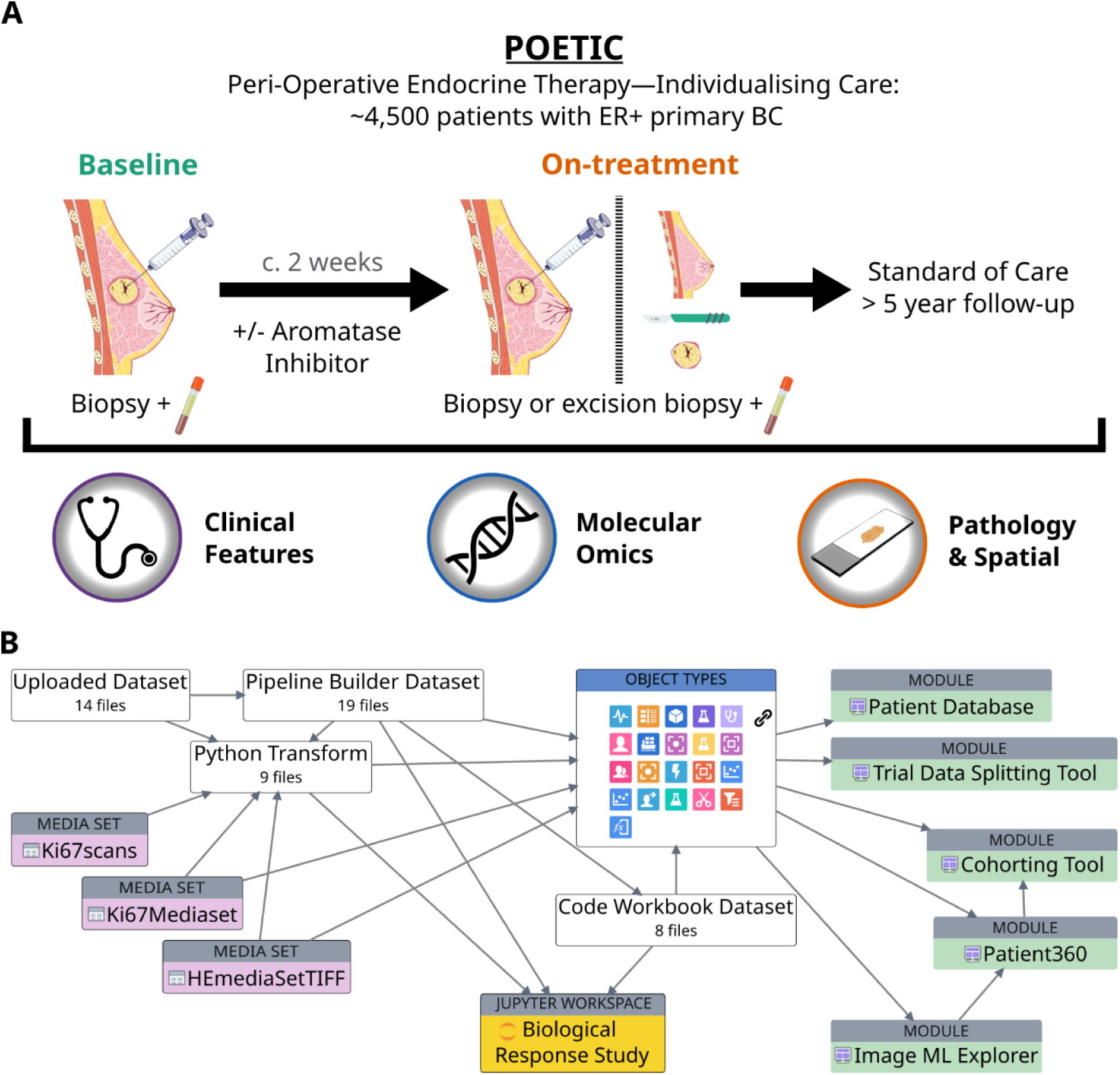
Overview of POETIC and the POETIC-AI ontology. **A.** Overview of the POETIC trial and data modalities generated from trial samples. **B.** POETIC-AI ontology showing relationships between clinical and molecular datasets, imaging and spatial media sets, analytical objects and application modules.

The paired baseline and surgical samples collected within POETIC have subsequently supported an extensive translational research programme. These samples are now characterised by bulk RNA sequencing, targeted mutational profiling, and quantitative proteomics, together with digitised haematoxylin and eosin (H&E) and Ki67 IHC, multiplex immunofluorescence, and spatial molecular profiling using the Geox Digital Spatial Profiler and the Cos Mx Spatial Molecular Imager. POETIC has therefore evolved from a clinical trial addressing a specific therapeutic question into a multimodal resource in which the biological feature changes resulting from a defined treatment perturbation can be examined across molecular, cellular, and tissue scales.

This evolution also changes how the trial data needs to be analysed. Individual modalities can be analysed independently, but doing so does not readily capture relationships between measurements made from the same patient, at different treatment timepoints, across different assays or within specific regions of the same tumour. These relationships become increasingly important as analyses move from individual biomarkers towards AI approaches that learn representations from high-dimensional data and integrate information across modalities. Furthermore, not all data modalities are inherently tabular and therefore easily interoperable; imaging and spatial datasets require data analysis workflows that first encode information into tabular formats. The analytical problem is thus not simply one of data volume or computational capacity, but of maintaining the structure of the clinical trial and its biospecimens throughout increasingly complex analytical workflows in trials like POETIC^3^.

For POETIC, this requires patients and treatment allocation to remain connected to samples, tissue sections, images, spatial regions, molecular measurements and subsequently derived features and data model outputs. It also requires analytical workflows ranging from conventional statistical analyses to image processing, spatial analysis and AI to operate within the same data environment. Importantly, because the data originates from a clinical trial, its integration must preserve provenance, reproducibility, security and appropriate control of sensitive information.

Here we present the POETIC Analytical Integration environment (POETIC-AI), a digital research environment designed to address these requirements. We use the commercial solution Foundry^4^ as a test case, implementing a graph-based ontology to represent the clinical and biological structure of POETIC and to connect patients, treatment allocation, specimens, timepoints, assays, images, spatial regions, analytical workflows, and derived measurements. We also introduce an approach for incorporating spatial omics images and their associated molecular data into Foundry, enabling these datasets to be linked to the corresponding tissue regions, specimens and clinical metadata within the same analytical environment. We describe how this environment supports integration and analysis across POETIC and use proliferation as a biologically grounded test case: reproducing the reduction in Ki67% following POAI and examining its heterogeneity; evaluating proliferation-associated signals across molecular, imaging and spatial modalities; and applying AI approaches to derive and connect features across these data types. Together, these analyses demonstrate how a clinical trial can be represented as a connected digital research environment for integrated multimodal and AI-based analysis.

## Results

### Multiscale and Multimodal Representation of POETIC Breast Cancer Clinical Trial

We constructed POETIC-AI as a digital representation of the clinical and biospecimen structure of POETIC, using a graph-based ontology to connect clinical, molecular, imaging and spatial data within a single governed analytical environment implemented using Foundry^4^ (Figure 1B).

To date, 14 datasets representing different data layers of POETIC have been incorporated into the environment. Relationships among patients, treatment allocation, sampling time points, specimens, assays, and derived molecular features were explicitly mapped within the ontology. Imaging and spatial data were incorporated as media sets and linked to their corresponding specimens and molecular data, while R- and Python-based analytical workflows and their derived outputs were retained within the same structure. This allowed analyses to operate on connected trial data while maintaining the relationship of each measurement to the patient, treatment, specimen and assay from which it was derived.

Five application modules were developed on this underlying data model: Patient Database, Clinical Trial Data Splitting Tool, Cohorting Tool, Image ML Explorer and Patient360. Together, these supported three main use cases: 1) definition of clinically annotated cohorts and generation of training and testing sets for downstream machine-learning analyses; 2) exploration of molecular profiles within newly or previously defined cohorts; and 3) linked visualisation and interrogation of pathology and spatial molecular data.

The resulting architecture provides a multiscale representation of POETIC in which data can be traversed from patient and treatment allocation through biospecimens to molecular assays, images, spatial regions and derived analytical features (Supplementary Figure 1A-D). This provides the underlying data structure for the cross-modal and AI analyses described below.

### Integrated Workflows Derive Orthogonal Measures of Tumour Proliferation

Ki67 was a predefined biomarker in POETIC, with Ki67% assessed from IHC at baseline and after two weeks of POAI providing a measure of tumour proliferation and early biological response to endocrine treatment^1,5^. We therefore used proliferation as a biologically grounded benchmark to determine whether different analytical workflows within POETIC-AI could recover the established treatment-associated phenotype across different data modalities.

We have implemented two complementary forms of analytical workflow. Python-based pipelines provided scalable processing of computationally intensive datasets, including whole-slide images, while graphical workflows incorporating R code enabled reproducible analyses directly linked to ontology-defined data objects. Using these approaches, we generated alternative measures of proliferation from digitised histopathology images and bulk gene expression (Figure 2A).

**Figure 2:**
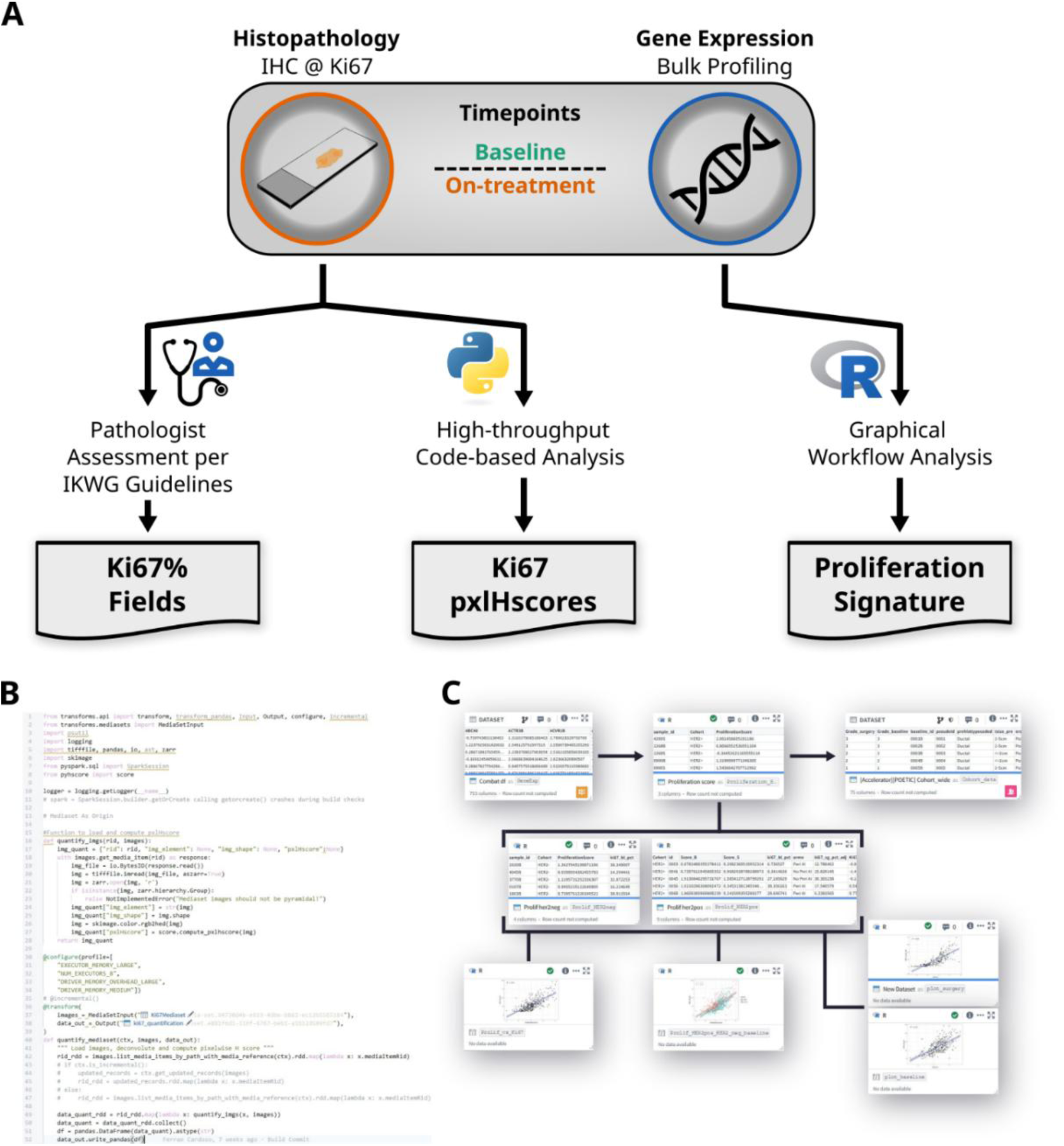
Derivation of proliferation metrics within POETIC-AI. **A.** Generation of three measures of proliferation: pathologist-assessed Kic7% fields, image-derived Kic7 pixel H-scores (pxlHscore) and gene expression-derived proliferation signature scores. **B.** Code-based image-analysis pipeline used to calculate Kic7 pxlHscores. **C.** Graphical workffow incorporating R analysis to calculate proliferation signature scores.

A code-based image-analysis pipeline quantified Ki67 staining from IHC whole-slide images to generate pixel-wise H-scores (Figure 2B)^6^. In parallel, a graphical workflow generated the clinically validated Prosigna® 18-gene proliferation signature^7^ from bulk gene expression data, including batch correction for technical variation between expression experiments (Figure 2C). These image- and transcriptome-derived measurements were then linked within the ontology to the corresponding patient, specimen and timepoint, together with the original expert field-based Ki67% assessment in POETIC trial. POETIC-AI therefore provided three orthogonal measures of proliferation derived from histopathologist assessment, digital image analysis and gene expression, allowing us to determine that the same treatment-associated biological signal could be observed across modalities.

### Treatment-associated Changes in Ki67% Reveal Increased Relative Field-level Heterogeneity

Given that a reduction in Ki67% after short-term aromatase inhibition is an established marker of biological response to endocrine therapy in POETIC^1,8^, we analysed the original field-level Ki67% measurements to reproduce this treatment-associated response and examined whether using multiple field values could additionally capture changes in intratumoral heterogeneity.

We evaluated ER+/HER2- and ER+/HER2+ bookend cohorts (See Methods) comprising 352 and 44 patients, respectively, selected to represent contrasting responses to POAI. Their Ki67% distributions in relation to the corresponding overall trial populations are shown in Figure 3A. The number of Ki67 fields assessed per sample was similar between the two cohorts and timepoints, with a mean of 5.1–5.7 fields (Figure 3B).

**Figure 3:**
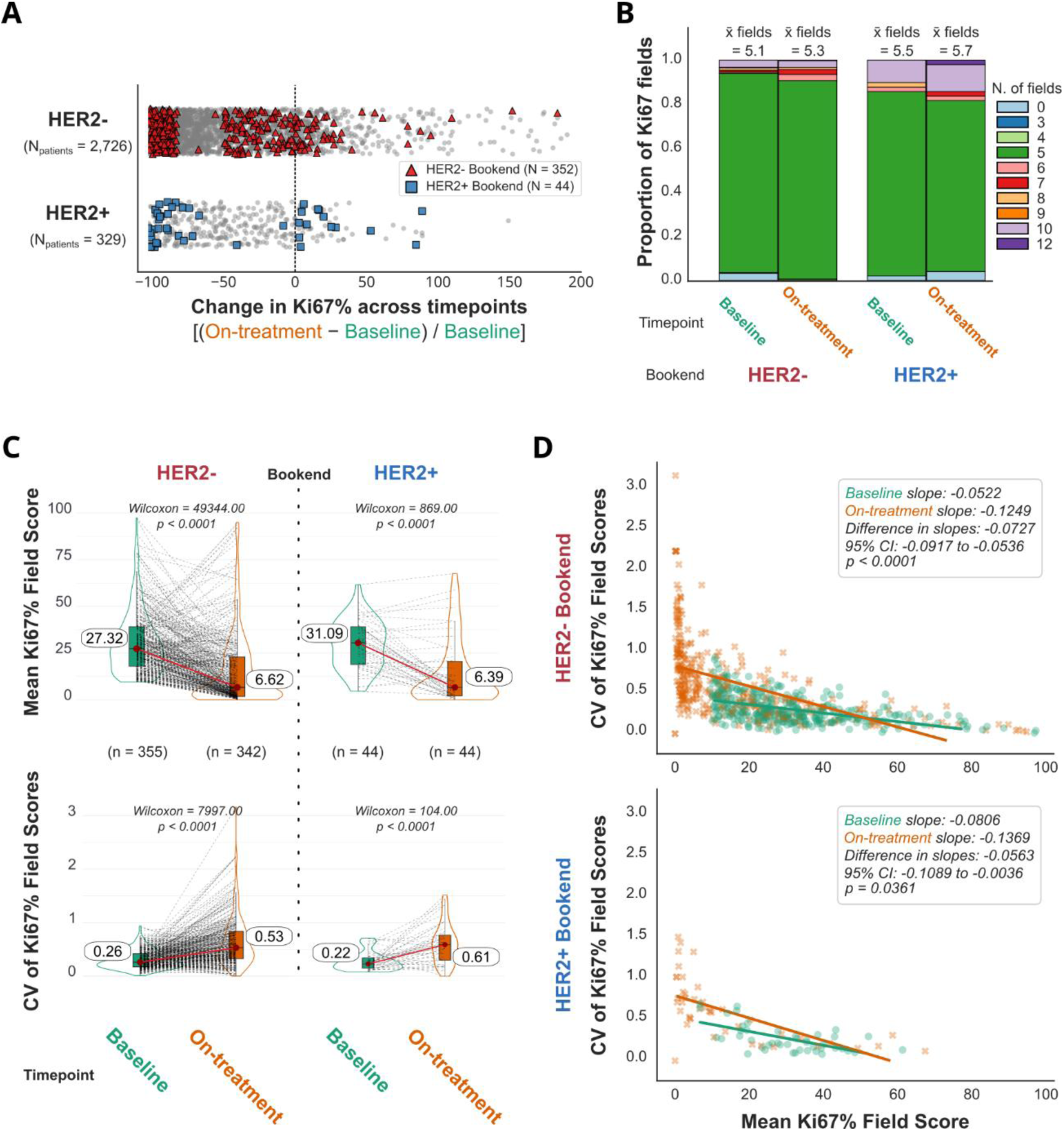
Changes in KiC7% and field-level dispersion following POAI. **A.** Changes in Kic7% distributions in HER2− and HER2+ POETIC cohorts, with bookend cases highlighted. **B.** Number of Kic7 fields assessed per sample in the HER2− and HER2+ bookend cohorts. **C.** Mean and coefficient of variation (CV) of Kic7% field scores at baseline and on-treatment. **D.** Relationship between mean Kic7% and field-level CV. Points represent samples from patients with complete baseline and on-treatment measurements. Lines show population-averaged estimates from cohort-specific Gaussian generalised estimating equation models accounting for repeated measurements within patients. Slopes represent the expected difference in Kic7 CV per 10-percentage-point higher mean Kic7.

Mean Ki67% values decreased between baseline and on-treatment samples in both cohorts. The relative dispersion of the Ki67 fields assessed, measured using the coefficient of variation (CV), increased from baseline to on-treatment samples (paired Wilcoxon tests, all P < 0.0001; Figure 3C), while absolute dispersion values decreased with the mean (Standard deviation, paired Wilcoxon tests, all P < 0.0001; Supplementary Figure 2A-B). Mean and CV values were comparable between the HER2- and HER2+ bookend cohorts (Supplementary Figure 2C-D).

Mean Ki67% values were inversely associated with relative field-level dispersion at both timepoints, with a stronger negative relationship at on-treatment than at baseline in both cohorts (Figure 3D). The on-treatment versus baseline difference in slope was −0.0727 in the ER+/HER2− cohort (95% CI −0.0917 to −0.0536; *P* < 0.0001) and −0.0563 per 10-percentage-point higher Ki67% in the ER+/HER2+ cohort (95% CI −0.1089 to −0.0036; *P* = 0.0361).

Since bookend cohorts do not represent intermediate responders, we repeated the analysis in the complete HER2+ POAI cohort (n=258 patients). We again observed a reduction in mean Ki67% values accompanied by increased relative field-level dispersion on treatment (Supplementary Figure 2E-F; paired Wilcoxon tests, P < 0.0001). Thus, the expected suppression of tumour proliferation following POAI was accompanied by increased relative dispersion of residual Ki67% across assessed tumour fields.

### Cross-modal Measurements Capture Different Aspects of Treatment-associated Proliferation

We next used the connected data structure to compare proliferation measured independently using gene expression, digital pathology and expert pathological assessment. We focused on the HER2+ bookend cohort, for which Ki67 IHC whole-slide images were available, providing 88 samples with all three proliferation measurements (Figure 4A).

**Figure 4:**
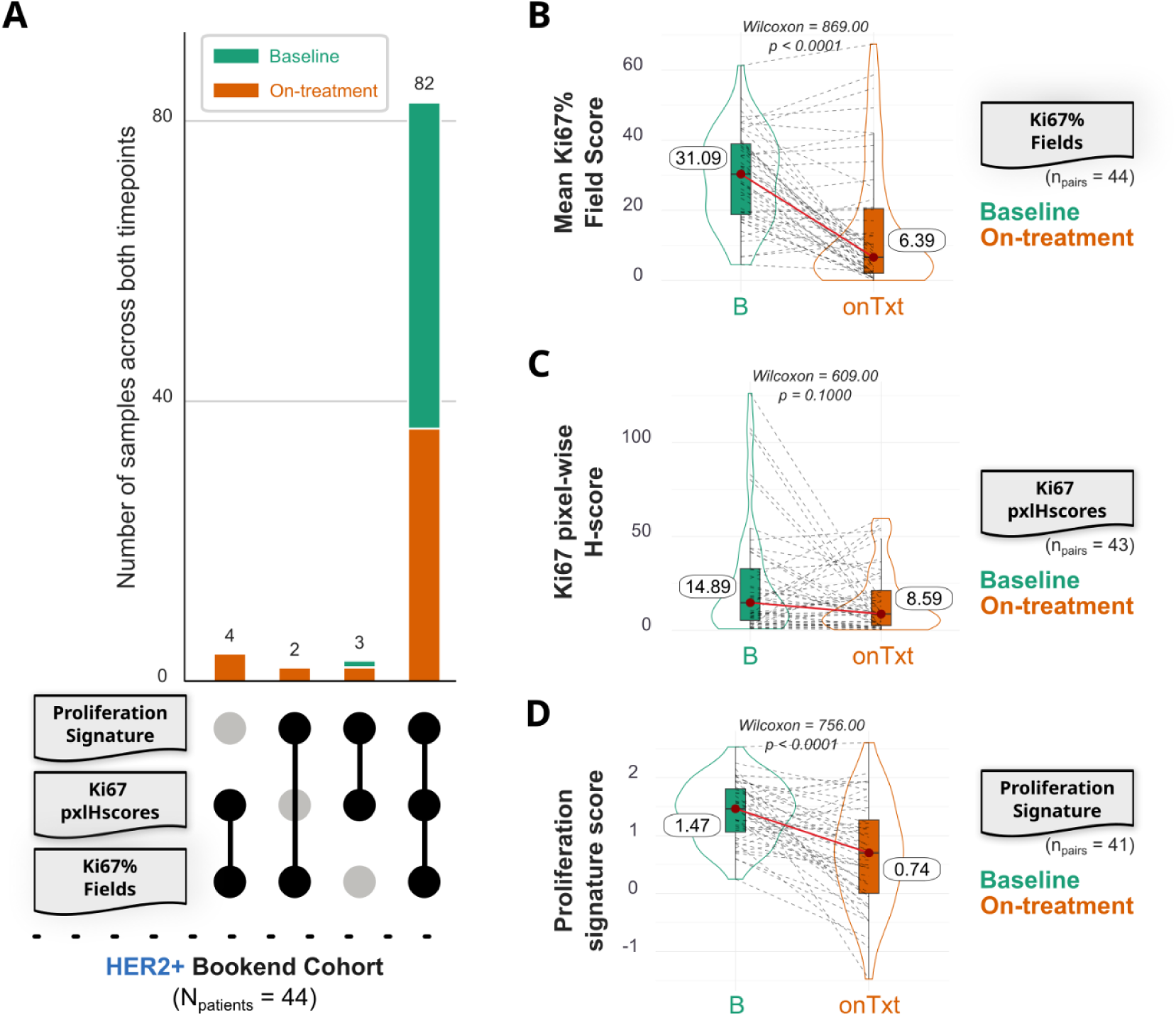
Cross-modal quantification of proliferation in the HER2+ bookend cohort. **A.** UpSet plot showing availability and overlap of the three proliferation measures. Forty-eight baseline and 40 on-treatment samples had all three measures available. **B-D.** Baseline and on-treatment proliferation measured using the gene expression proliferation signature (**B**), Kic7 pxlHscore (**C**) and mean pathologist-assessed Kic7% (**D**).

Both pathologist-assessed Ki67% and the gene expression proliferation signature showed marked reductions following POAI (paired Wilcoxon tests, both P < 0.0001; Figure 4B and 4D). In contrast, the image-derived Ki67 pxlHscore did not show a significant treatment-associated reduction (P = 0.15; Figure 4C).

Despite being generated from different modalities, all three measurements were positively correlated. The strongest association was observed between pathologist-assessed Ki67% and the gene expression proliferation signature (Spearman *r* = 0.834). The image-derived pxlHscore showed more modest correlations with pathologist-assessed Ki67% (*r*=0.472) and the proliferation signature (*r*=0.327) (Table 1).

**Table 1:** Spearman correlation coefficients among the three proliferation quantification modalities.95% confidence intervals and p-values were also stated in the table.

|  | Ki67% Fields | Proliferation Signature | Ki67 pxlHscore |
| --- | --- | --- | --- |
| Ki67% Fields | 1 | 0.828 (0.750 – 0.884)<br>$p < 0.0001$ | 0.461 (0.283 – 0.609)<br>$p < 0.0001$ |
| Proliferation Signature | | 1 | 0.316 (0.118 – 0.490)<br>$p = 0.0023$ |
| Ki67 pxlHscore |  |  | 1 |

These analyses showed that measurements designed to represent the same biological phenotype were not necessarily equivalent across modalities. The strong correlation between the gene expression proliferation signature and pathologist-assessed Ki67% indicated that the treatment-associated proliferation signal could be recovered from an orthogonal molecular modality. In contrast, the simpler pixel-based quantification of Ki67 staining showed weaker cross-modal correlations and did not reproduce the treatment-associated reduction observed with expert assessment. This suggests that quantifying staining intensity alone does not capture all the biological information incorporated into expert assessment of tumour proliferation, providing a benchmark against which more context-aware digital pathology and AI approaches can be evaluated.

### Linking Multimodal Measurements to Their Morphological and Spatial Context

We next extended the Foundry solution to allow findings identified through cohort-level and computational analyses to be traced back to the tissue from which they were derived. Whole-slide pathology and spatial-omics data were therefore incorporated into the ontology and connected to their corresponding patients, specimens, molecular measurements and spatial annotations.

The H&E viewer enables morphological assessment of tumour architecture and tissue composition (Figure 5A). The Ki67 viewer links individual field-level assessments to their whole slide images, providing an overview of the IHC staining accompanied by the field-level assessments from expert pathologists (Figure 5B), which allows variation in proliferation identified computationally to be examined directly within its morphological context.

**Figure 5:**
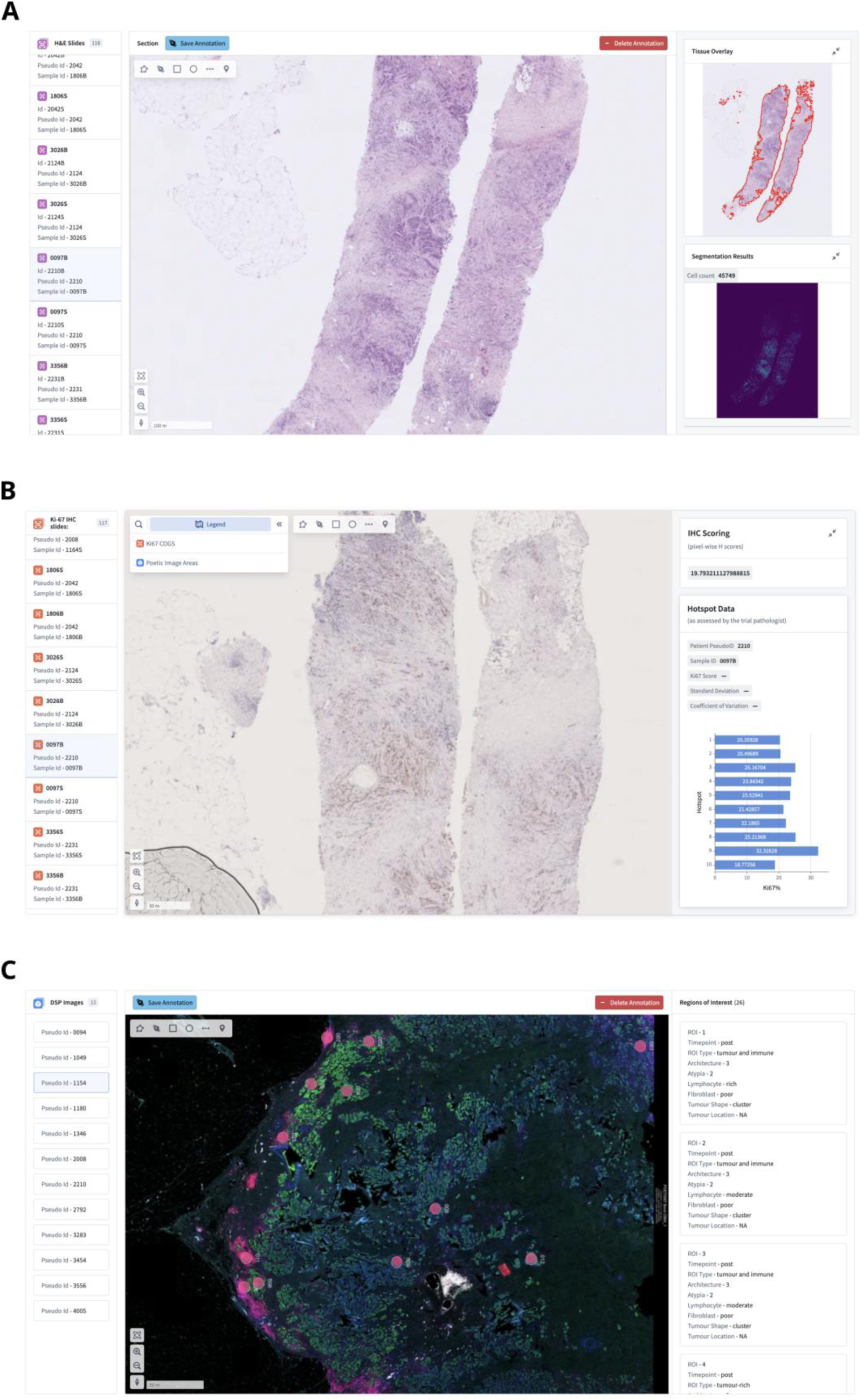
Integrated exploration of pathology and spatial molecular data. **A.** H&E whole-slide image viewer. **B.** Kic7 image viewer linking field-level Kic7 assessments to their corresponding locations within the whole-slide image. **C.** Spatial molecular viewer linking GeoMx regions of interest to immunoffuorescence images and spatial transcriptomic measurements

For spatial transcriptomic data, GeoMx immunofluorescence images and regions of interest (ROIs) were linked to their corresponding molecular profiles (Figure 5C). This allows investigators to move from a patient or molecular phenotype identified through cohort-level analysis to the corresponding tissue region and examine molecular measurements in relation to tumour morphology and the surrounding microenvironment.

The same structure also provides a route for AI-derived features and predictions to be returned to their underlying clinical and tissue context. Rather than treating model outputs as independent analytical endpoints, these can be linked to the patient, treatment timepoint, specimen, image or spatial region from which they were generated and subsequently compared with measurements from other modalities. In this way, POETIC-AI supports analysis at scale while retaining the clinical, morphological and spatial context required for biological interpretation of multimodal and AI-derived findings.

## Discussion

In this study, we developed POETIC-AI, a digital research environment linking the clinical, molecular, imaging, and spatial data generated through the POETIC trial. Using proliferation as a biologically grounded test case, we reproduced the established reduction in proliferation following POAI, examined the field-level variation underlying conventional Ki67 measurements, compared proliferation across different data modalities, and linked these measurements back to their morphological and spatial context. More broadly, we demonstrated how these data can be represented and analysed as a connected, computable research resource rather than as a collection of independent datasets.

This challenge is increasingly recognised across biomedical research. The FAIR (Findability, Accessibility, Interoperability, and Reusability) principles^9^ and initiatives such as the NCI Cancer Research Data Commons (CRDC, https://datacommons.cancer.gov/) have established frameworks for making multimodal biomedical data and analytical workflows accessible, interoperable, reusable and available within secure computational environments. POETIC-AI applies these principles to a complementary challenge: preserving the structure of a longitudinal, biospecimen-rich clinical trial as new molecular, imaging, spatial and computational data are generated. Its ontology links these measurements to patients, treatment allocation, sampling timepoints, specimens and clinical outcomes, maintaining their context and provenance as the resource evolves. However, an ontology is only as informative as its underlying data and metadata, and integrating datasets generated over many years, technologies and research projects required substantial biological knowledge and multidisciplinary curation. Developing and maintaining such environments should therefore be considered part of the long-term scientific stewardship of a clinical trial, rather than solely an informatics task.

POETIC-AI supports both code-based and graphical analytical workflows, and we consider both important for translational research as they provide complementary routes for bespoke method development and reuse of established analyses by researchers with different computational expertise. Because both operate on the same trial representation, derived measurements can be returned to the ontology with their analytical provenance and reused in subsequent studies rather than remaining as isolated project outputs. As the number of computationally derived biomarkers and AI-generated features increases, maintaining the provenance of how each measurement was generated will become increasingly important.

We used Ki67 as our initial biological test case because the primary POETIC analysis established the prognostic importance of on-treatment proliferation following short-term endocrine therapy^1^. By retaining the individual Ki67 field assessments rather than only their sample-level averages, we reproduced the expected reduction in proliferation while also examining variation between fields. Mean Ki67% decreased following POAI, whereas relative field-level dispersion increased in both ER+/HER2+ and ER+/HER2− bookend cohorts, perhaps suggesting increased intra-tumour heterogeneity in response to endocrine therapy exposure. A similar pattern was observed in the wider POAI-treated HER2+ population, suggesting that this observation was not restricted to patients selected for extreme response.

These findings illustrate how reducing multiple measurements to a single biomarker value can remove potentially informative biological variation. Two tumours with a similar mean on-treatment Ki67% could, for example, represent either relatively uniform suppression of proliferation or a tumour containing predominantly suppressed regions alongside residual areas of higher proliferation. Whether these represent biologically or clinically distinct states remains unknown. Our analysis was not designed to establish spatial heterogeneity, as the Ki67% fields were not selected using a systematic spatial sampling strategy. In addition, the coefficient of variation depends on the mean and therefore requires cautious interpretation when the mean Ki67% falls substantially following treatment. Direct spatial mapping and ultimately association with clinical outcome will be required to determine whether residual heterogeneity provides information beyond conventional on-treatment Ki67%.

Pathologist-assessed Ki67% and the 18-gene proliferation signature derived from genes in the Prosigna® assay were strongly correlated and both captured the expected reduction following POAI, showing that the treatment-associated phenotype could be recovered across histopathological and molecular modalities. However, the measurements are not interchangeable: Ki67% is restricted to morphologically identified tumour cells, whereas the transcriptomic signature is an aggregate signal influenced by tumour cellularity, sampling, and tissue composition. This distinction was clearer for the image-derived pixel-wise H-score^6^, which showed weaker correlations with both measurements and a directionally similar, but statistically non-significant, reduction following POAI. This demonstrates that converting a pathology image into a quantitative measurement does not necessarily capture the tumour recognition, tissue architecture, and biological context incorporated into expert assessment. More sophisticated computational pathology approaches could address these limitations by incorporating tumour detection, cellular morphology, tissue architecture, and learned image features. POETIC provides a relevant benchmark for developing such methods because its paired samples allow models to be evaluated not only by agreement with existing biomarkers, but also by their ability to capture treatment-induced biological change and potentially identify additional features of response.

Linking computational findings to their source tissue enables multimodal measurements to be interpreted within their morphological and spatial context. In POETIC-AI, Ki67 field measurements and spatial molecular profiles remain connected to their corresponding whole-slide images and annotated GeoMx regions, allowing differences between modalities to be investigated in relation to tumour content, morphology, sampling location, and the surrounding microenvironment. To our knowledge, this represents the first reported implementation within Foundry or similar platforms to integrate and interactively explore spatial-omics data alongside their source images, spatial annotations, molecular measurements, and associated clinical trial data. The same framework preserves the provenance of computationally derived features, supporting their biological interpretation within the patients, specimens, and tissue regions from which they originated.

Several limitations should be considered. The response-enriched cohorts used for some analyses do not capture the full spectrum of endocrine response within POETIC, and the relatively small HER2+ cohort limited the statistical power of some comparisons. These findings therefore require evaluation in the wider POETIC population and, where appropriate, in independent datasets. POETIC-AI has also been implemented in a single clinical trial, and the generalisability of its ontology and applications to other trial designs remains to be demonstrated. Foundry provided the governance, ontology, computational and application-development capabilities required to develop and test this implementation, but the underlying trial-centric data model and analytical approach are not intended to depend on a particular commercial platform. That said, their portability remains untested and deployment in other environments would require adaptation to local infrastructure. A longer-term model for multimodal clinical-trial research may therefore rely on common data structures and portable analytical components that can operate across appropriately governed environments.

In conclusion, POETIC-AI provides one implementation of a trial-centric multimodal research environment. Using proliferation as a benchmark, we demonstrate how established trial biomarkers can be revisited at greater resolution, compared across modalities and returned to their tissue context. As clinical trials become increasingly multimodal and AI-enabled, maintaining these connections will be important if complex molecular and computational measurements are to be translated into biologically interpretable, reproducible and clinically testable biomarkers.

## Methods

### Data Handling and Analysis Platform

Data for the POETIC-AI study was uploaded to the Foundry platform as 14 individual raw datasets. The disparate data sources were all harmonised in platform using ontology-based data pipeline operations to the same primary key; a patient POETIC pseudo-ID for patient-level observations, and a composite pseudo-ID appended with a sample indicator (B or S) for timepoint-based observations (i.e. baseline and on-treatment timepoints). The raw datasets for image-based modalities had to be processed in-platform for conversion into mediaset objects; supporting thumbnail previews and lazy-loading on various applications developed within POETIC-AI.

The 5 distinct modules and derived applications were developed using in platform tooling and linked to their relevant input and output objects (e.g. saved annotation polygons in the Image ML Explorer) in the ontology.

### POETIC Cohort Descriptions

#### POETIC ER+/HER2- Bookend Cohort

From the POETIC POAI treatment arm, HER2- breast cancer tumours with baseline Ki67 IHC > 10% were selected. Poor responders (PRs) to POAI were defined as the patients in the lowest 15% of anti-proliferative response to POAI based on the proportional change in Ki67 between baseline and on-treatment timepoints^11^. PRs were matched on baseline Ki67 categories (10-20%, 20-30%, and ≥30%) with good responders (GRs) (defined as the top 50% of anti-proliferative response) to ensure that the GRs were selected not biased for low proliferation tumours^11^. In total, RNA-seq data was available for 341 (174 GRs and 167 PRs) paired sample sets^11^.

#### POETIC ER+/HER2+ Bookend Cohort

From the POETIC POAI treatment arm, we selected 46 patients with HER2+ breast cancer as the bookend cohort. We selected these cases based on their anti-proliferative response to POAI, defined as the proportional change in Ki67 between baseline and on-treatment timepoints. The cohort comprised 15 tumours in the top 15% of Ki67 reduction among the ER+/HER2+ POAI-treated population, classified as good responders (GR), and 16 tumours in the bottom 15%, classified as poor responders (PR). We additionally included 14 tumours showing an intermediate Ki67 reduction but classified as HER2- enriched (HER2-E) at baseline; these tumours were analysed as a separate HER2-E GR group. Their inclusion was based on prior molecular analyses showing that the HER2-E subtype at pretreatment is associated with early resistance to POAI^12^. One additional tumour was included as a special case within the PR group: although its Ki67 reduction fell just outside the bottom 15% threshold, it showed an atypical biological response characterised by subtype switching from Luminal A to HER2-E after POAI. NanoString Breast Cancer 360 data and digitised Ki67 IHC-staining images were available for all patients in this cohort.

### Transcriptomics Proliferation Signature and Scoring

Transcriptomics data was acquired from both RNA-sequencing and NanoString Breast Cancer 360 codeset. Batch correction was performed in R (version 3.6), between the two gene expression experiments, using the “ComBat” function from the sva package (automatically pulling the latest version from Bioconductor).

The gene expression-based proliferation signature score was calculated as the mean expression value of 18 proliferation genes: *ANLN, CCNE1, CDC20, CDC6, CDCA1, CENPF, CEP55, EXO1, KIF2C, KNTC2, MELK, MKIc7, ORCc, PTTG1, RRM2, TYMS, UBE2C,* and *UBE2T*^7^.

### Immunohistochemistry Quantification

Ki67 IHC scans were processed in platform using a custom computational pipeline to compute a variant of the pixel-wise H-score as implemented in the pyhscore package^10^. Default intensity thresholding values were used for the calculation (0.05 for haematoxylin, 0.12, 0.24, and 0.6 for low, medium and high DAB respectively), which is defined as follows:

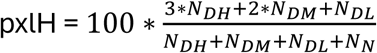

where:

- *N_DH_* is the number of pixels with high DAB intensity,
- *N_DM_* is the number of pixels with medium DAB intensity,
- *N_DL_* is the number of pixels with low DAB intensity,
- *N_N_* is the number of pixels with positive haematoxylin but negative DAB intensity.

A mediaset ontology object containing reduced resolution images was generated from the raw images (3^rd^ largest resolution level of 20X scans) to reduce the computational requirements of the pipeline. This pipeline was deployed on the platform’s infrastructure using a custom spark profile that distributes the images to be processed in parallel across 8 executor cores with a large memory setting, owing to the need to transform from RGB into HED colour spaces in memory. Results were then saved to Foundry’s ontology as a dataset by the Spark driver node.

### Comparison of Proliferation Metrics

Wilcoxon tests were used to compare the proliferation metrics between pre-treatment and on-treatment, evaluating measurements from both timepoints as paired observations per patient. Mann-Whitney tests were performed to compare the proliferation metrics between HER2- and HER2+ cohorts as unpaired independent sets.

Wilcoxon and Mann-Whitney tests were performed in R using the ggstatsplot package (v0.13.5)

To evaluate the relationship between mean Ki67 and relative field-level dispersion, cohort-specific Gaussian generalized estimating equation (GEE) models with an exchangeable correlation structure were fitted separately to the ER+/HER2+ and ER+/HER2− bookend cohorts using the statsmodel Python package (v0.14.6). Models included mean Ki67 (expressed per 10 percentage points), timepoint, and a Ki67-by-timepoint interaction term, with patient identifiers used to account for repeated measurements. The interaction term was used to assess whether the association between mean Ki67 and relative field-level dispersion differed between pre-treatment and on-treatment samples. Analyses were restricted to patients with complete paired measurements at both timepoints. Spearman correlation tests were performed to evaluate the correlations between the proliferation quantification metrics regardless of the timepoint. The 95% confidence intervals of the Spearman’s *γ* were calculated as 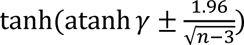, where n is the number of data points in the correlation test.

### Digital Pathology and Spatial Image Visualisation

Digital pathology and spatial images are inherently large, often comprising gigapixel-scale datasets that can be challenging to render and visualise efficiently. To address this, we stored the large image files as mediasets containing a thumbnail image alongside multiple pyramidal resolution levels. This multiresolution architecture enables the image viewer to dynamically select the appropriate image level according to the user’s zoom state, providing rapid rendering while minimising computational and storage demands.

For a selection of HCE scans we also deployed a pipeline for tissue detection and cell detection in order to populate the additional layers in the Image ML Explorer app. Tissue detection was performed using intensity thresholding on the RGB colour space of the image, with the threshold automatically selected using Otsu’s implementation in the skimage *Python* package. Cellular segmentation was performed using the cellpose *Python* package with the cyto2 segmentation model^13^.

Fluorescent images from GeoMx and cellular and biological details on selected ROIs were uploaded as raw data into platform. These images were then converted in platform into the annotation-supporting Geotiff format. To link the ROI details with the images, ROI annotation overlays were generated in platform and then programmatically linked to the ROI details using a JSON-like data structure, which could then be used by the Image ML Explorer. Similarly, platform-native tools and data pipelines were used to link the Ki67 IHC scans to the corresponding Ki67 field spot data.

### Figure assembly and plotting tools

Where relevant, individual figure panels were first generated in R with packages ggplot2 and ggstatsplot, or in Python with UpSetPlot, seaborn and matplotlib. The graph ontology overview panels were exported from the platform as SVGs images. Final editing and figure assembly was performed using the open-source vector graphics editor Inkscape and using open assets from the NIH BioArt and Bioicons repositories.

## Data Availability

All data produced in the present study are available upon reasonable request to the authors and subject to the relevant approval process.

## Acknowledgments

POETIC is co-sponsored by The Institute of Cancer Research and The Royal Marsden NHS Foundation Trust. POETIC is funded by Cancer Research UK (CRUK/07/015) and coordinated by the ICR-CTSU. ICR-CTSU also receives programme grant funding from Cancer Research UK (grant number C1491–A15955).

We acknowledge NHS funding to the NIHR Biomedical Research Centre at The Royal Marsden and The Institute of Cancer Research. The views expressed are those of the authors and not necessarily those of the NIHR or the Department of Health and Social Care. We thank all the POETIC participants and staff for their help and assistance. We thank also Breast Cancer Now and CRUK for additional funding. We also thank the past and present colleagues on the POETIC Trial Management Group, and the POETIC Independent Data Monitoring Committee and Trial Steering Committee.

Finally, we thank Justin Whatling, Philip Lindstedt, Nicholas McQuibban, Arjun Banerjee, and Linda Zhao for providing technical support on the Foundry platform.

