## Supplemental Figures for "A Digital Environment for Multi-modal Relational Management, Representation, and Analysis of Cancer Clinical Studies"

### Supplementary Figures

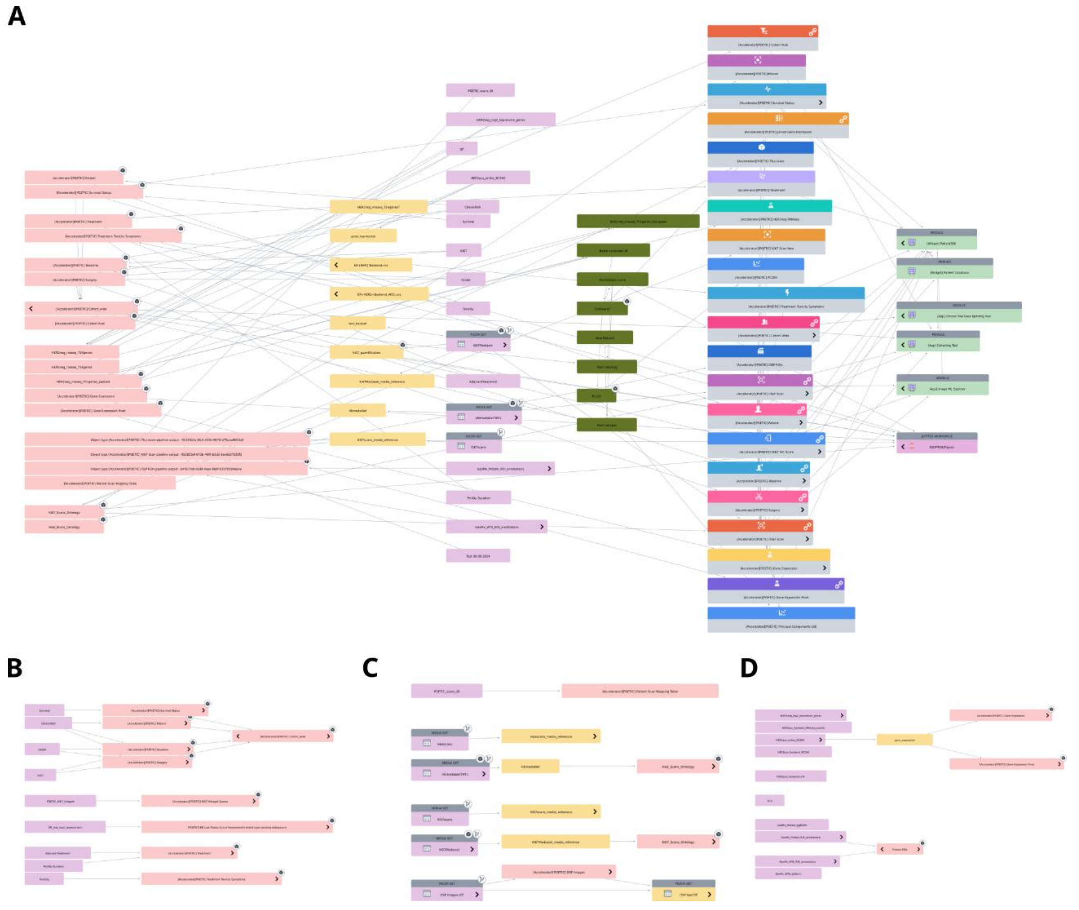


***Supplementary Figure 1***. *A. Main ontology objects and relationships. B-D. Ontology subset to (B) clinical datasets, (C) imaging and spatial datasets, and (D) molecular assays, respectively.*

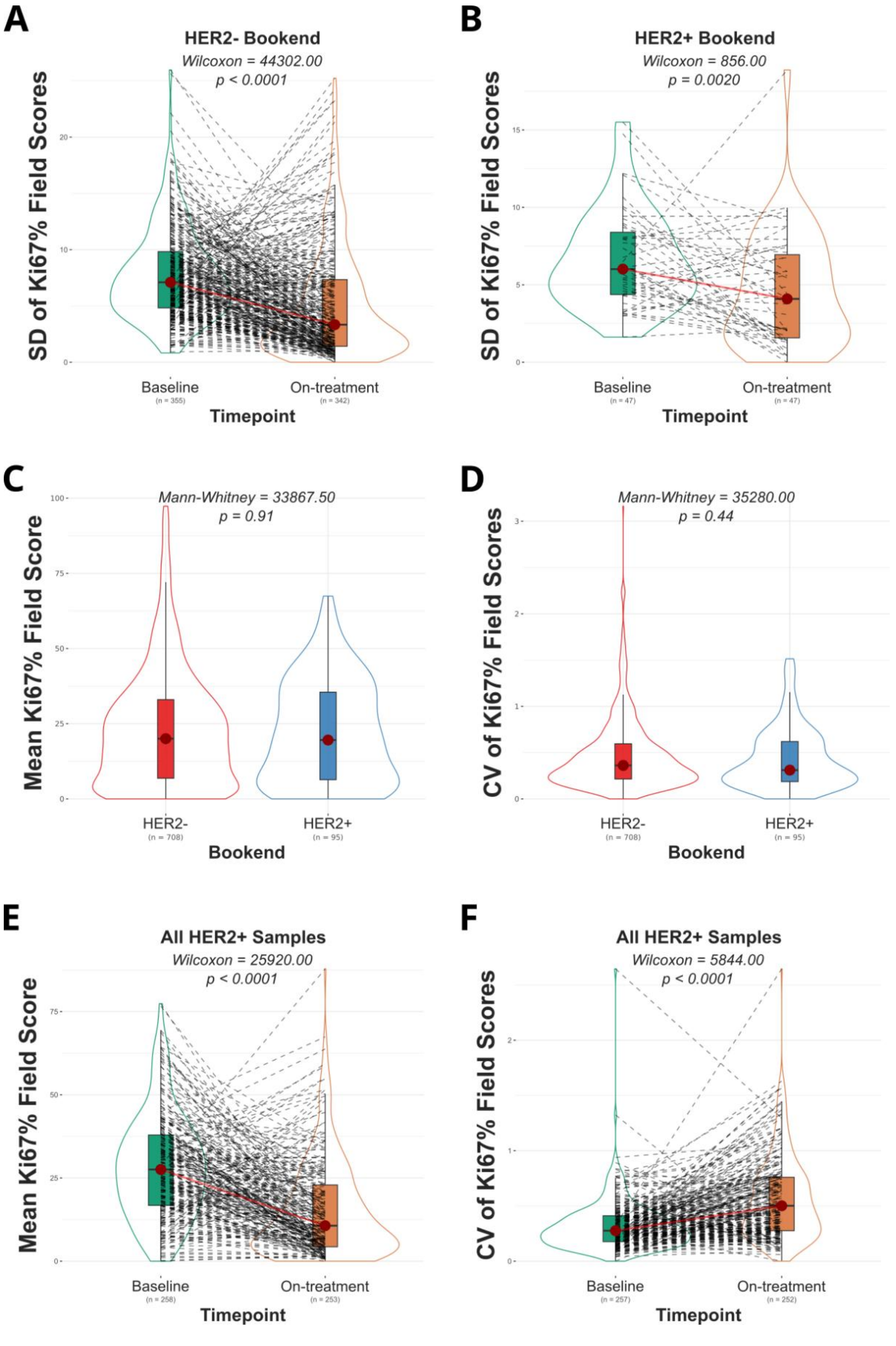


***Supplementary Figure 2***. *A. Mean Ki67% distribution between HER2- (n=708) and HER2+ (n=95) bookend cohorts, regardless of timepoint. B. Ki67 heterogeneity (coefficient of variation) distribution between HER2- (n=708) and HER2+ (n=95) bookend cohorts, regardless of timepoint. C. Mean Ki67% distribution between HER2+ POAI group (n=258) baseline and on-treatment timepoints. Wilcoxon test was performed to evaluate the differences between the two timepoints. D. Ki67 heterogeneity (coefficient of variation) distribution between HER2+ POAI group (n=258) baseline and on-treatment timepoints. Wilcoxon test was performed to evaluate the differences between the two timepoints.*
